# Functional respiratory capacity in the elderly after COVID-19 – a pilot study

**DOI:** 10.1101/2021.09.03.21263076

**Authors:** Filipe Alexandre Pereira, Maria Teresa Tomás

## Abstract

**Background:** The pandemic spread of SARS-CoV-2 has led to an unprecedented outbreak of viral pneumonia. Despite the current focus of worldwide research being the characterization of post-COVID-19 sequelae, the level of functional impact that this disease causes in the elderly who have presented moderate, severe or critical manifestations is still unknown.

**Objective:** To identify the main consequences/sequelae on functional respiratory capacity in the elderly after CoViD-19.

**Methodology:** A cross-sectional study was carried out in the community. Functional aerobic capacity (2min step test), dyspnea perception (modified Medical Research Council Dyspnea Questionnaire), respiratory and peripheral muscle strength (maximum inspiratory and expiratory pressure, grip strength) and the Frailty Index (Clinical Fragility Scale) were assessed in 25 community-dwelling individuals aged ≥65 years, who have had a diagnosis of CoViD-19 for up to 6 months, and in an equal number of elderly people with the same characteristics without a known diagnosis of CoViD-19.

**Results:** The elderly with a diagnosis of CoViD-19 up to 6 months presented a decrease in the values of maximum inspiratory pressure (p=0.001) and maximum expiratory pressure (p=0.015), in aerobic capacity (p<0.001) with significant presence of desaturation on exertion (p<0.001), and increased values of dyspnea perception (p=0.001) and Frailty Index (p=0.026).

**Conclusion:** Significant changes were found in the functional respiratory capacity of elderly patients diagnosed with CoViD-19 for up to 6 months, when compared with elderly individuals without a known diagnosis of CoViD-19. It is not possible to extrapolate the results obtained to the Portuguese population, however these results may be an important indicator in the characterization of sequelae in the elderly after infection by SARS-CoV-2.

## INTRODUCTION

The year 2020 was marked by the pandemic spread of a new coronavirus, called *Severe Acute Respiratory Syndrome-Coronavirus-2* (SARS-CoV-2)^1^, which causes *Coronavirus Disease – 2019* (CoViD-19). Symptomatic presentation is characterized by mild respiratory infections or pneumonia of moderate to severe intensity. Pneumonia can cause hypoxemic respiratory failure due to bilateral interstitial infiltration with severe changes in the ventilation-perfusion ratio, pulmonary shunt, lung parenchyma necrosis and pulmonary fibrosis caused by consolidation of pulmonary exudate, which may progress to *Acute Respiratory Distress Syndrome* (ARDS) and multisystemic failure^2^.

By the end of June 2021, more than 880000 confirmed cases had been registered in Portugal, with a mortality rate exceeding 17100 deaths. Worldwide, more than 181 million cases are confirmed in 191 countries/regions, with mortality exceeding 4 million deaths^3^.

Overall, approximately 80% of all CoViD-19 patients, confirmed by laboratory testing, have mild to moderate disease, with or without pneumonia, 13.8% have severe disease (dyspnea, respiratory rate ≥30/minute, peripheral blood oxygen saturation (SpO2) ≤93%, partial pressure of oxygen/inspired oxygen fraction <300, pulmonary infiltration >50% of the lung field in 24-48 hours) and 6.1% has critical illness (respiratory failure, septic shock and multisystem failure)^4^. Consequently, thousands of patients worldwide are hospitalized for acute care, where between 75-80% have prolonged hospitalizations (±21 days)^5^. In Portugal, a 15% hospitalization rate was reported, with an ICU admission rate of 1.5% and a mortality rate of 2.5%^6^.

However, CoViD-19 doesn’t affect only the lungs, being currently described as a multisystemic disease^7^. Simultaneously, a hyper inflammatory state caused by the patient’s immune response (cytokine storm) seems to be responsible for the dysfunction increase in various organs and systems^8^, being associated with cardiac^9^, neurological^10^ and musculoskeletal complications^11^.

As this is a disease with a short evolution, it’s not clear whether it will leave permanent pulmonary or physical sequelae and to what extent, existing a need for studies to characterize the sequelae and their evolution. Lung tissue alterations such as ground glass opacities, consolidation, vascular thickening, bronchiectasis, pleural effusion, mosaic paving pattern and presence of nodules may exist in more than 80% of survivors^12^. A study^13^ suggests that pulmonary fibrosis will become one of the main sequelae, showing that 45% of patients presented signs of pulmonary fibrosis one month after infection and developed pulmonary fibrosis 3 to 6 months after infection.

The set of symptoms that can remain 12 or more weeks after the initial infection was called *Long COVID*, with reports of severe symptoms persistence for more than 6 months after the initial infection, even in moderate initial manifestations. Research on *Long COVID* is growing, and there’s already initial evidence^14^ of 55 long-term complications associated with CoViD-19, highlighting for clinical practice the decrease in SpO2 on exertion, thromboembolic disease, myocarditis or pericarditis, dysautonomia, pulmonary interstitial disease, myelopathy, neuropathy and neurocognitive disorders, with a study^15^ pointing to a prevalence that can reach 10% of cases with persistent symptoms. However, there’s still no consensus in the literature on the definition of *Long COVID*, its cause, prevalence, susceptibility, severity, diagnosis or treatment^14^.

Although some aspects of this disease remain uncertain, there seems to be no doubt that elderly people, potentially fragile and subject to more comorbidities, are at greater risk of severe and/or fatal manifestation^16^, which is also described in the Portuguese population^6^. This is probably due to the fact that the elderly have a more vulnerable immune system and less physiological reserve in face of an aggressor event, as well as a greater probability of comorbidities, which makes them more exposed and more likely to develop this disease^17^. An investigation^18^ involving frailty and hospitalization indicate that this condition is a better evolution and negative outcome predictor in situations of disease, than age or comorbidities, and this relation is also described in CoViD-19^19^. It’s currently predicted that this disease could result in significant morbidity for 3-6 months, with 45% of patients in need of medical and social assistance and 4% of patients in need of inpatient rehabilitation, with consequent pressure on medical and rehabilitation services beyond 12 months^7^.

Considering the worldwide impact caused by CoViD-19, understanding this disease functional repercussions in recovered patients is of great interest. Despite the current research focus being the characterization of the sequelae, the functional impact and the potential consequences that this disease causes in the elderly who have presented SARS-CoV-2 infection is still unknown.

Taking this into account, the aim of this study was to identify possible sequelae in the functional respiratory capacity in elderly people who were diagnosed with CoViD-19, asymptomatic and with moderate, severe or critical symptoms. Therefore, functional aerobic capacity, respiratory and peripheral muscle strength, levels of dyspnea and Frailty Index were evaluated in a community-dwelling group of individuals over 65 years of age, diagnosed with CoViD-19 up to 6 months before the start date of the study, in comparison with an equal number of elderly people with the same characteristics without a previous diagnosis of CoViD-19.

## METHODS

This was a quantitative study with a cross-sectional observational research design, with a control group. The population were individuals who were diagnosed with SARS-CoV-2 infection within 6 months, asymptomatic or with moderate, severe or critical symptoms. The study sample consisted of community-dwelling individuals aged ≥65 years, diagnosed with CoViD-19 within a period of up to 6 months before the evaluation carried out in this study, who voluntarily agreed to participate after signing a informed consent by themselves or their health caregiver. Individuals who refused to sign an informed consent (self or caregiver) were excluded. The control group consisted of community-dwelling individuals, with identical age and gender characteristics but without a diagnosis of CoViD-19, which agreed to participate in the study. The informed consent was signed by all participants, and the study was approved by the Ethics Committee of ESTeSL.

Data collection took place in Lisboa e Vale do Tejo area with 50 participants evaluated. Sample general characterization data such as age; gender; Body Mass Index (BMI); comorbidities; occurrence, type and duration of hospitalization, were collected. The variables under study were: functional aerobic capacity; peripheral and respiratory muscle strength, dyspnea perception and Frailty Index.

Functional aerobic capacity was assessed with the 2-Minute Step Test (TMST), used in different studies to assess aerobic capacity in the elderly^20^, which is a valid and reliable test^21^, recommended by the *American Physical Therapy Association*^22^ to assess aerobic exercise capacity in patients with and after CoViD-19. During this test, SpO2 was monitored using finger oximetry (mindray PM-60 oximeter) in order to measure desaturation on exertion existence, as recommended by the *American Thoracic Society/European Respiratory Society*^23^ guidelines in the assessment of patients after CoViD-19. A decrease in SpO2 ≥4% was considered clinically significant^24^.

The perception of dyspnea was assessed using the modified Medical Research Council Dyspnea Questionnaire (mMRC), according to the recommendations of the *Portuguese Association of Physiotherapists and Interest Group in Cardio-Respiratory Physiotherapy* (GIFCR) in the evaluation of patients with CoViD-19^25^.

Peripheral muscle strength was assessed by measuring isometric grip strength (kg) with the JAMAR® hydraulic dynamometer. This measurement is valid and suitable not only for the assessment of upper limb muscle strength^26^, but it’s also associated with general peripheral muscle strength^27^, which shows great importance due to its relation with the individuals’ functional capacity, allowing the determination of risk levels for future incapacity^28^.

Respiratory muscle strength was assessed by measuring maximal inspiratory pressure (MIP) and maximal expiratory pressure (MEP), as recommended by the GIFCR in the evaluation of patients with CoViD-19^25^, using a MicroRPM® manovacuometer. As recommended by the guidelines^29^, the use of non-invasive manovacuometers to measure maximal respiratory pressures (cm H2O) quantifies respiratory muscle strength in various populations with different characteristics, showing excellent reliability.

The Frailty Index was identified through the Clinical Frailty Scale (CFS), a valid and reliable instrument in the assessment of frailty that, in addition to being easy and quick to apply, is a predictor of institutionalization and mortality^30^.

The data obtained were analysed using the *Statistical Package for Social Sciences – IBM SPSS 25*.*0* software. T test for 2 independent samples and Mann-Whitney test were used to group comparison in the different variables. The level of statistical significance chosen was p<0.05.

## RESULTS

The study sample consisted of 50 individuals (28 men and 22 women) older than 65 years (70.2 ± 5.6 years). 25 individuals constituted the post-covid group (after diagnosis of CoViD-19 up to 6 months) and 25 individuals the control group (without CoViD-19 diagnosis) (Table 1). The assessment in the post-covid group occurred 4.5 ± 0.6 months after diagnosis of CoViD-19.

**Table 1.** Sample characterization data: sex, age and Body Mass Index

|  |  | Post-covid |  | Control |  | p value |
| --- | --- | --- | --- | --- | --- | --- |
|  |  | Frequencies | Mean + SD | Frequencies | Mean + SD |  |
| Sex | Female | 12 (54.5%) |  | 10 (45.5%) |  | p=0.396 |
|  | Male | 13 (46.4%) |  | 15 (53.6%) |  |  |
| Age |  |  | 69.6 ± 6.0 |  | 70.8 ± 5.1 | p=0.149 |
| Body Mass Index |  |  | 28 ± 4.7 |  | 27.2 ± 4.4 | p=0.832 |

In the study sample, the participants presented heart disease and metabolic disease as the main comorbidities (Table 2) and only 4% presented no comorbidity. In the post-covid group, 20% of participants had 1 comorbidity, 36% 2 comorbidities and 44% 3 or more comorbidities, while in the control group 44% had 1 comorbidity, 32% 2 comorbidities and 16% 3 or more comorbidities. In the post-covid group only 2 were asymptomatic, 12 were hospitalized (20.9 ± 7.2 days) and of these, 5 were hospitalized in the Intensive Care Unit (36 ± 6.5 days) (Table 3).

**Table 2.** Sample characterization data: comorbidities

|  | Post-covid | Control | p value |
| --- | --- | --- | --- |
| Heart disease | 18 | 16 | p=0.554 |
| Respiratory disease | 8 | 4 | p=0.193 |
| Cancer disease | 2 | 4 | p=0.394 |
| Osteoarticular disease | 10 | 7 | p=0.381 |
| Psychiatric illness | 2 | 0 | p=0.155 |
| Vascular/lymphatic disease | 3 | 1 | p=0.307 |
| Metabolic disease | 12 | 8 | p=0.257 |
| Neurological disease | 2 | 1 | p=0.561 |
| Obesity | 3 | 3 | p=1 |

**Table 3.**
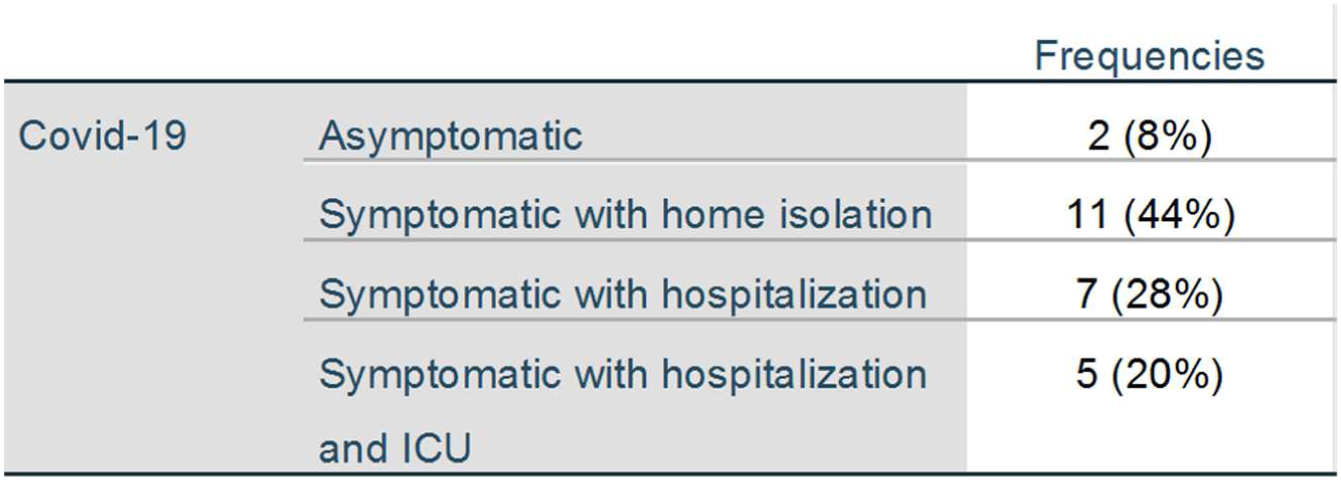
Sample Characterization Data: CoViD-19 pattern

|  |  | Frequencies |
| --- | --- | --- |
| Covid-19 | Asymptomatic | 2 (8%) |
|  | Symptomatic with home isolation | 11 (44%) |
|  | Symptomatic with hospitalization | 7 (28%) |
|  | Symptomatic with hospitalization and ICU | 5 (20%) |

### Frailty

There were differences registered between the two groups for the Frailty Index (p=0.026), with higher levels of frailty in the post-covid group compared to the control group (table 4).

**Table 4.**
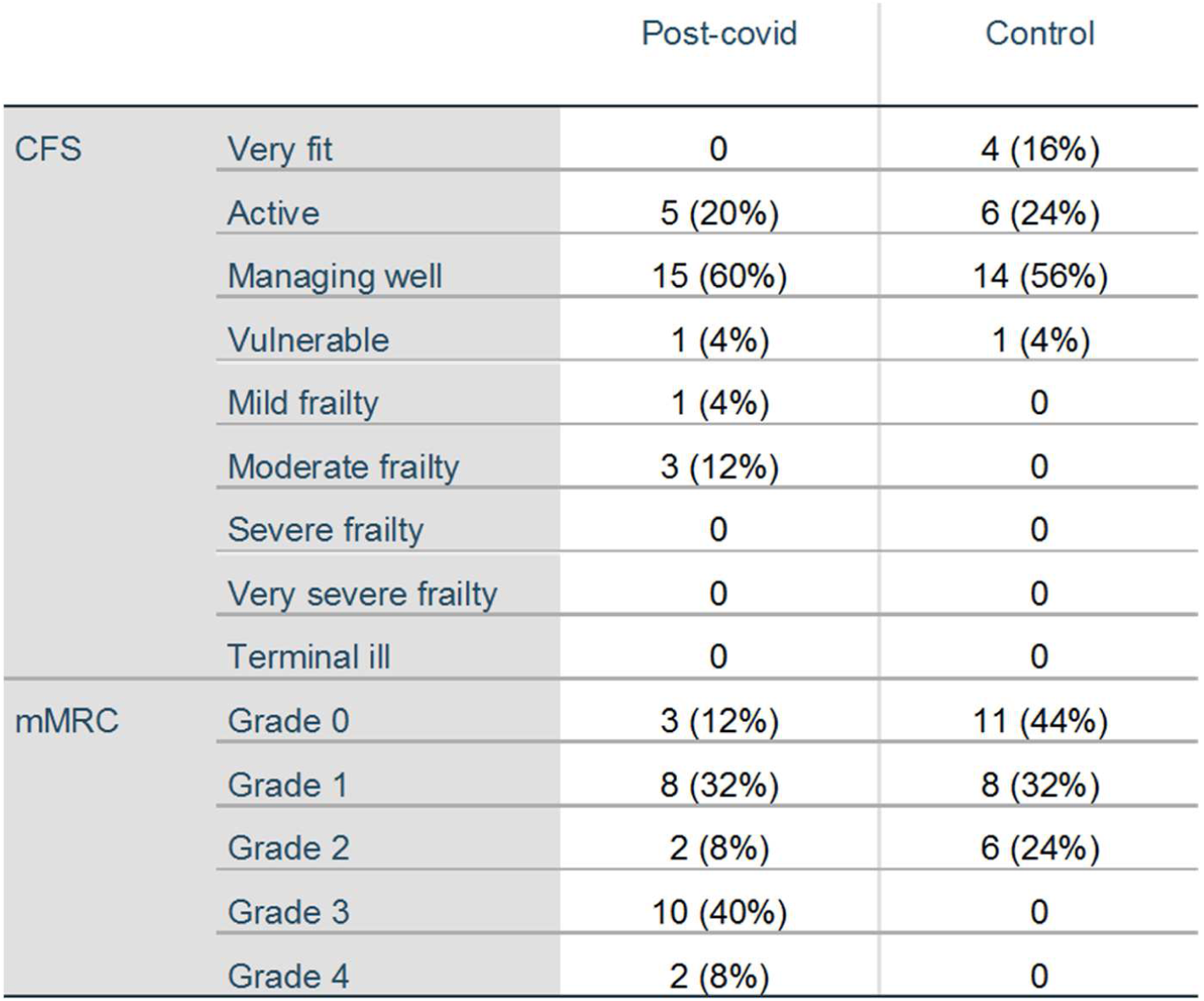
Results: Clinical Frailty Scale (CFS) and modified Medical Research Council Questionnaire (mMRC)

### Dyspnea perception

The post-covid group showed differences in relation to the control group (p=0.001) in the mMRC classification, with higher mean levels of dyspnea perception than the levels perceived by the individuals in the control group (table 4).

### Respiratory muscle strength

The study groups presented different values, with the control group presenting higher values in relation to the post-covid group in both MIP (p=0.001) and MEP (p=0.015), indicating a muscular strength decrease of the respiratory muscles in the post-covid group compared to the control group (Table 5).

**Table 5.** Mean, standard deviation and p value results: maximal inspiratory pressure (MIP), maximal expiratory pressure (MEP), grip strength and 2minute step test (TMST)

|  | Post-covid<br>Mean + SD | Control<br>Mean + SD | p value |
| --- | --- | --- | --- |
| MIP | 55.6 ± 28.7 | 82.8 ± 24.8 | **p=0.001** |
| MEP | 87 ± 28.4 | 105.2 ± 22.5 | **p=0.015** |
| Right grip strength | 28.1 ± 9.5 | 32 ± 9.0 | p=0.135 |
| Left grip strength | 26.5 ± 9,3 | 30.5 ± 8.3 | p=0.118 |
| TMST | 40.8 ± 15.4 | 74.2 ± 21.2 | **p<0.001** |

### Peripheral muscle strength

There were no differences between the two groups in grip strength values, both in the right hand (p=0.135) and in the left hand (p=118) (Table 5).

### Functional aerobic capacity

Regarding the TMST, the two groups showed different results, with the post-covid group registering lower values (p<0.001), demonstrating a lower functional aerobic capacity (Table 5). During the TMST performance, differences were observed between the groups concerning desaturation on exertion (p<0.001). In addition to the post-covid group having presented higher desaturation values, it was in this group that almost all clinically significant desaturation (≥4%) was registered, which occurred in only 1 participant in the control group (table 6).

**Table 6.** Results: Desaturation on exertion

|  |  | Post-covid | Control |
| --- | --- | --- | --- |
| Desaturation on exertion | 0 | 2 (8%) | 18 (72%) |
|  | 1 | 2 (8%) | 4 (16%) |
|  | 2 | 6 (24%) | 1 (4%) |
|  | 3 | 3 (12%) | 1 (4%) |
|  | 4 | **6 (24%)** | **1 (4%)** |
|  | 5 | **2 (8%)** | 0 |
|  | 6 | 0 | 0 |
|  | 7 | **1 (4%)** | 0 |
|  | 8 | 0 | 0 |
|  | 9 | **1 (4%)** | 0 |
|  | 10 | **2 (8%)** | 0 |

## DISCUSSION

Although *Long COVID* characterization being the current focus of investigation, to our knowledge there are no studies comparing functional variables among elderly people diagnosed with CoViD-19 vs without a diagnosis of CoViD-19. However, given the heterogeneity of CoViD-19 clinical presentation, it’s essential to have simple tools to assess and monitor the impact of symptoms on the respiratory system and on these patients functionality, the main outcomes of physical therapy intervention in patients with CoViD-19 sequelae.

The variables and measurement instruments used in this study are recommended by the existing guidelines^23^ for post-covid patients assessment in sequelae characterization, and have been used to compare physical therapy interventions in different contexts^8,37,39^. The variables that characterize the study sample, namely advanced age, gender, BMI, comorbidities and hospitalization length seem to be among the most significant for CoViD-19, in line with other investigations^31,33^ that correlate these variables with the increased risk of hospitalization and functional capacity of post-covid patients after hospital discharge. The absence of significant difference between the post-covid group and the control group in the sample characterization variables under study, including age (p=0.148), gender (p=0.396), BMI (p=0.832) and comorbidities (p’s >0.05), allows its comparisons.

Investigations concerning post-covid sequelae exist mostly in patients who required hospitalization. However, it seems to be important to characterize the sequelae by the pattern of disease registered, as done in the present study, where 44% of patients evaluated in the post-covid group belonged to the symptomatic with home isolation subgroup. A longitudinal study^32^ described the presence of symptoms, including dyspnea and fatigue, up to 7 months after the initial infection in patients with mild and moderate disease, concluding that these sequelae may occur in the long term even in patients with mild and moderate disease, reinforcing the need of sequelae characterization in this subgroup. However, the balance registered in the sample’s distribution, between 52% of non-hospitalized participants and 48% of hospitalized participants, is noteworthy.

Regarding the Frailty Index, the difference verified in the frailty levels between the two groups in our study, is in agreement with the results obtained by Owen and colleagues (2021)^33^, which correlated the Frailty Index, through the CFS, and mortality among hospitalized patients with and without a diagnosis of CoViD-19, demonstrating that there are also differences in the Frailty Index between the groups, with a higher percentage of frailty in the CoViD-19 group.

Grip strength, one of the main determinants of frailty and sarcopenia, as well as a predictor of dysfunction, morbidity and mortality, has been associated with CoViD-19. It’s strongly correlated with respiratory muscle strength (MIP and MEP) and both are inversely proportional to age^34^, with descriptions^35^ of this relationship in healthy populations. A larger study^31^ points out that grip strength assessed by a dynamometer is inversely correlated with risk of hospitalization for CoViD-19, assuming grip strength as an independent risk factor for the severity of CoViD-19. However, it can’t be said that the post-covid group has lower grip strength as there was no statistically significant difference. This finding is supported by the fact that the values obtained in the post-covid group doesn’t present significant differences in relation to normative values^36^. The fact that a significant percentage (52%) of the sample wasn’t hospitalized may have contributed to this result.

Based on existing scientific evidence from the pulmonary sequelae study in individuals who have recovered from other CoV outbreaks (SARS-CoV and MERS-CoV), as well as the existing evidence on SARS-CoV-2^37^ infection, testing respiratory and gas exchange are extremely important, including MIP and MEP assessment^38^. MIP is a strong indicator of diaphragm muscle strength, while MEP measures the strength of the abdominal and intercostal muscles, particularly important for effective coughing and related to dyspnea. Decreased expiratory muscle strength can lead to air trapping, while decreased inspiratory muscles can lead to atelectasis. Several reasons for post-covid respiratory muscle weakness have been suggested, including viral damage-induced myositis, muscle wasting and deconditioning due to prolonged lodging, myopathy caused by corticosteroids, and critical illness-associated polyneuropathy or myopathy^38^. The restrictive pattern observed in several investigations^37^ on lung function in patients after CoViD-19 may be partially due to respiratory muscle weakness, as reported by a follow-up study^39^ at 3 and 6 months after hospital discharge, where there was a persistent decrease in MIP and MEP, and by another investigation^40^ on respiratory function in the post-covid convalescence phase, which points to an 80% decrease in the predictive values of MIP and MEP, respectively. These investigations are in line with the results obtained in our study, were was registered a significant difference in the measurement of maximal respiratory pressures between the post-covid group and the control group. This finding is supported by the fact that the post-covid group presents measurements at 67% and 60% of the predictive values^29^ of MIP and MEP, respectively.

The decrease in respiratory muscle strength partially contributes to the observed changes in aerobic capacity. The significant decrease in aerobic capacity measured by TMST with oxygen desaturation during exercise verified in this study, is in line with the results obtained in aerobic capacity assessment, in rehabilitation programs for patients with CoViD-19 after hospital discharge^41^. This finding is supported by the fact that the values recorded in the post-covid group were below the mean of predictive values^21^, specifically between 45% and 58% in men and between 41% and 66% in women.

Desaturation on exertion is one of the main sequelae observed after SARS-CoV2^24^ infection. The results obtained clearly demonstrate the existence of this sequel, with significant desaturation in 48% of post-covid participants compared to the control group, where significant desaturation occurred in 1 participant (4%), who had previous oncologic respiratory disease. This can be explained by evidence^37^ indicating that the distinctive feature of CoViD-19 is extensive damage to alveolar epithelial cells and endothelial cells with secondary fibroproliferation, indicating a potential for chronic alveolar vascular and alveolar remodelling, leading to pulmonary fibrosis and/or pulmonary hypertension and mainly compromising the diffusion-perfusion capacity, as reported by this meta-analysis^37^.

Although this study limitation (small sample), the characterization of the sequelae found is essential in the elderly age group. Decreased respiratory muscle strength and aerobic capacity can affect the ability to perform ADL, in addition to implying an increase in the vulnerability of the immune system as well as a reduction in physiological reserve in face of a potential aggressor event. This has consequences in the increase of frailty, which is directly related to the decrease in functionality, institutionalization and mortality. To our knowledge, there were no studies specifically carried out with older adults (>65 years of age). Taking into account the burden on social and rehabilitation services, the early inclusion of these patients in pulmonary and/or cardiovascular rehabilitation programs seems very important, aiming for the recovering of their respiratory and functional capacity.

## CONCLUSION

In the study sample, it was found that elderly people diagnosed with CoViD-19 up to 6 months, showed changes in functional respiratory capacity, namely decreased respiratory muscle strength and functional aerobic capacity with desaturation on exertion, and consequently an increase in the subjective perception of dyspnea, with higher rates of frailty, compared to an identical group without a previous diagnosis of CoViD-19.

Research studies on post-covid sequelae characterization is necessary, with larger samples representative of the population, and which also include patients with mild and moderate symptoms, as there are indications that these may also be prevalent significant. The knowledge of these sequelae is important to being able to act earlier in the recovery of the verified limitations, which can represent a very high impact on the quality of life in elderly patients.

## Data Availability

Data collected available in SPSS statistics output document

